# Association of Sex and Other Demographic Characteristics with Hemoglobin A1c in Older Adults with Type 1 Diabetes: Cross-sectional analysis of the U.S. Premier Healthcare Database

**DOI:** 10.1101/2025.11.13.25340150

**Authors:** Trisha Nagulagari, Wendy Novicoff, Genevieve R. Lyons, Maria C. Sanchez Valenzuela, Nicolas A. Reyes, Sue A. Brown, Kaitlin M. Love

**Affiliations:** Division of Endocrinology and Metabolism, Department of Medicine, University of Virginia Health, Charlottesville, VA; Department of Public Health Sciences and Orthopedic Surgery, University of Virginia, Charlottesville, VA; Department of Public Health Sciences, University of Virginia, Charlottesville, VA; Division of Endocrinology and Metabolism, Mercy Hospital Fort Smith, Fort Smith, AR; Division of Endocrinology, Diabetes, and Metabolism, University of Miami, Miami, FL; AdventHealth Translational Research Institute, Orlando, FL

## Abstract

**Objective:** In type 1 diabetes, results for hemoglobin A1c (HbA1c) sex-based differences are mixed with limited data in later ages. The study objective was to determine if female sex is independently associated with higher HbA1c in the United States older adult population.

**Research Design and Methods:** A cross-sectional analysis was performed on real world Premier Healthcare data from January 2020–December 2022 examining adults with type 1 diabetes, ages 55-75. Multivariate linear and logistic regression models were used to determine demographic variables independently associated with HbA1c. Predictor variables included age, sex, race, ethnicity, insurance payor, testing location, marital status, atherosclerotic cardiovascular disease.

**Results:** Of 12,088 individuals with HbA1c data (52.4% women, mean age 64.7 ± 5.8 years), HbA1c was 0.13% (95% CI: [0.08, 0.18], *P*<0.001) higher on average in females and 0.81-0.97% higher in Black patients adjusting for all other variables. Female sex (unadjusted odds ratio 1.13, 95% CI: [1.05, 1.21], *P*<0.001, adjusted odds ratio [aOR] 1.16, 95% CI: [1.07, 1.26], *P*<0.001), Black compared to White race (aOR 1.78, 95% CI: [1.49, 2.12], *P*<0.001), and Medicaid compared to Managed Care payor (aOR 1.63, 95% CI: [1.28, 2.07], *P*<0.001) were independently associated with increased odds of HbA1c above target, ≥7% for ages 55-64 and ≥8.5% for ages ≥ 65.

**Conclusions:** In this cohort with type 1 diabetes, females experienced slightly higher HbA1c and higher odds of HbA1c above recommendations. Individuals with Black race and Medicaid insurance payor experienced clinically significantly higher HbA1c levels. Further interventions to address HbA1c disparities are needed.

**Twitter summary:** Hemoglobin A1c was slightly higher for women, ages 55 to 75, with type 1 diabetes in a real world healthcare database. Major racial and insurance based hemoglobin A1c disparities were observed.

**Article Highlights:**

- Why did we undertake this study?

- Limited, mixed data exists examining hemoglobin A1c (HbA1c) sex-differences in people with type 1 diabetes in later ages.
- What is the specific question(s) we wanted to answer?

- In older age groups, do women have higher HbA1c levels, adjusting for demographic characteristics which influence HbA1c?
- What did we find?

- HbA1c was 0.13% higher in women compared to men adjusting for other variables. Women had higher odds of HbA1c above age-based targets.
- Black race was associated with a 0.81% and 0.97% higher HbA1c compared to White and Asian races respectively.
- What are the implications of our findings?

- Mechanistic study is needed to understand higher HbA1c in women at these age groups, and further intervention is needed to reduce health disparities.

---

Type 1 diabetes accelerates cardiovascular disease (CVD) death by 10-15 years (1,2). Disparities in healthcare exist for people with type 1 diabetes whereby certain vulnerable groups experience excess CVD risk. This is particularly observed in groups with lower income and less educational attainment (3). Individuals in minority racial groups experience a markedly higher burden of CVD (4,5). Gender disparities are also present in CVD risk. Presence of type 1 diabetes exaggerates cardiovascular risk for young adult women, essentially outweighing the cardiovascular protection conferred by female sex (6,7). In fact, women with type 1 diabetes experience a mortality risk that is greater than men without diabetes (6). Type 1 diabetes confers greater excess cardiovascular mortality risk for women compared to men (6,7). It is possible that suboptimal risk factor mitigation exacerbates these disparities. We recently observed, in a real-world electronic healthcare record (EHR) study of the Premier database examining a United States (U.S.) population of older adults with type 1 diabetes, women are less likely than men to be treated to recommended low density lipoprotein cholesterol thresholds (8).

In type 1 diabetes, hemoglobin A1c (HbA1c) lowering is critical to mitigate diabetes related complications including cardiovascular mortality (9,10). Data examining sex-based differences in HbA1c optimization in adult populations are mixed with some studies showing higher HbA1c in women (11–13) and others showing no difference (14,15). The disparate results could be due to the age groups studied as most studies detecting a difference had a higher mean age, over 40 years. A recent study from the Canadian National Diabetes Repository detected a HbA1c separation by sex with women having a higher HbA1c around the time of the menopausal transition, which was not statistically significant with the relatively small sample size (n = 296) (16). This mirrored lived experience reported by women with type 1 diabetes involved in that study (16). Additionally, while the U.S. has many diverse type 1 diabetes observational cohort studies there is limited real world data in the U.S. type 1 diabetes population and this may limit generalizability. Thus, additional U.S. studies which incorporate data from real-world settings are important to address these potential gaps.

The objective of this study was to determine if female sex is independently associated with elevated HbA1c, adjusting for other demographic factors known to impact HbA1c, in a U.S. cohort of older adults with type 1 diabetes. We set out to examine if the following predictor variables are associated with HbA1c: age, sex, race, ethnicity, insurance payor, marital status, testing location, atherosclerotic CVD (ASCVD). Our *a priori* hypothesis was that female sex, racial and ethnic minority groups, and Medicaid insurance would be associated with increased HbA1c adjusting for all other variables. We predicted this given cardiovascular risk disparities in these populations and our recent findings concerning for suboptimal treatment of cardiovascular risk factors in women and individuals with Medicaid insurance coverage in this age group (8).

## Research Design and Methods

We conducted a cross-sectional cohort analysis of older adults with International Classification of Disease (ICD)-10 diagnostic code for type 1 diabetes (E10.*) included in the Premier Healthcare Database (PHD) as previously reported (8). PHD is a U.S. hospital system-based, real world electronic medical record database including more than 315 million patients from 1,322 health care systems. The study was granted Human Subjects Research Institutional Review Board exemption by the University of Virginia (submission #302573).

### Eligibility

<u>Inclusion:</u> individuals with ICD-10 diagnostic code for type 1 diabetes, aged 55 to 75 years with outpatient location encounters across multiple specialties between January 1, 2020 to December 31, 2022 having HbA1c values were included. We selected the age range 55-75 years to include patients at high cardiovascular risk and avoid potential influence of pregnancy on glycemic targets. <u>Exclusion:</u> we excluded individuals with prolonged inpatient or emergency department encounters to limit the confounding impact of stress or infection mediated hyperglycemia. Individuals in palliative care clinical settings were excluded due to impact on treatment targets.

### Variables

The outcome variable was HbA1c reported in %. HbA1c values recorded in SI units (mmol/mol) or implausible values (i.e. 1, 33, 2010, 12866, 62249) consistent with data entry error were excluded. If duplicate HbA1c values were reported for a single participant, the lowest value was selected. Only 17 duplicate values were recorded spanning more than 1 week duration and resulted in a clinically significant classification change of HbA1c <8.5% in 2 cases and <7% in 3 cases. Predictor variables included age, sex, race, ethnicity, insurance payor (commercial, Managed care, Medicaid, Medicare/other non-Medicaid government insurance, other [including Worker’s compensation, Direct Employer compensation, or other], or uninsured [self-pay, indigent]), marital status, clinical setting (clinic, diagnostic evaluation for blood work or imaging alone, home health/rehabilitation, observation hospitalization [24-hour admission or less], outpatient surgery, infusion/hemodialysis, other), and cardiovascular disease diagnosis. Clinical setting was included due to the presence of patients in a pre-surgical setting represented in this cohort, which could confound outcomes. For example, patients receiving surgery could be required or personally motivated to achieve a lower HbA1c to improve surgical outcomes. Cardiovascular disease diagnosis included ICD-10 codes for ischemic coronary artery disease (I25.*), cerebral infarction (I63.*) and peripheral vascular disease (I73.8).

### Statistical analysis

Using SAS 9.4 (SAS Institute, Cary, NC), a multivariate linear regression model was used to determine which of the predictor variables were uniquely associated with HbA1c after adjusting for other predictor variables. To assess differential effect of HbA1c level across age by sex, we assessed for interaction between sex and age. To assess whether female sex was associated with higher HbA1c in certain racial groups, we assessed for interaction between sex and race. To determine if ASCVD diagnosis influenced HbA1c level by sex, we assessed for interaction between sex and ASCVD diagnosis. Additionally, to confirm whether the multivariate regression provided clinically relevant information about HbA1c levels above target, a logistic regression model was used to determine which of the predictor variables were uniquely associated with elevated hemoglobin A1c after adjusting for all other predictor variables. Elevated HbA1c was defined as ≥7% for individuals ages 55 to 64 years and ≥8.5% for individuals ages 65 years and older based on American Diabetes Association Guideline Recommendations during a majority of the observation period (17–19). Threshold for statistical significance was set at p ≤ 0.01. We selected this more stringent p-value to reduce likelihood that the significant results occurred by chance in light of the large sample size and number of predictor variables examined.

### Data and Resource Availability

Data used in this study are available from Premier Healthcare Solutions, Inc. Restrictions apply to the availability of this data, which were used under license for the current study and therefore are non-transferable in accordance with a data use agreement with Premier Healthcare Solutions, Inc.. Analytic code used in the current study are available from the corresponding author upon reasonable request.

Patient and Public Involvement: Patients or the public were not involved in the design, conduct, reporting, or dissemination plans of this research.

## Results

Of the 173,178 outpatients in the PHD cohort with a diagnosis code for type 1 diabetes, 12,088 unique patients had HbA1c data collected in an outpatient environment in this database (Consort diagram presented in Supplemental Figure S1). The small portion of cohort with outpatient HbA1c data could relate to the predominately inpatient nature of the PHD database or the observation period occurring during the COVID-19 pandemic. Demographic characteristics are presented in **Table 1**. Mean age was 64.7 ± 5.8 years. Of the 12,088 individuals, 6330 (52.4%) were females and 331 (2.7%) individuals identified as Hispanic ethnicity and 784 (6.5%) as Black race. HbA1c was tested predominately (59.8%) in a laboratory/imaging only encounter. Histogram of HbA1c values is presented in **Figure 1A**. Mean HbA1c was 7.7 ± 1.5%. Mean HbA1c across the cohort age distribution, separated by sex is shown in **Figure 1B**, and HbA1c scatter plot by sex is shown in **Figure 1C**. On average, HbA1c was higher in younger ages and lower in older ages with women having higher HbA1c compared to men until age 67.

**Figure 1:**
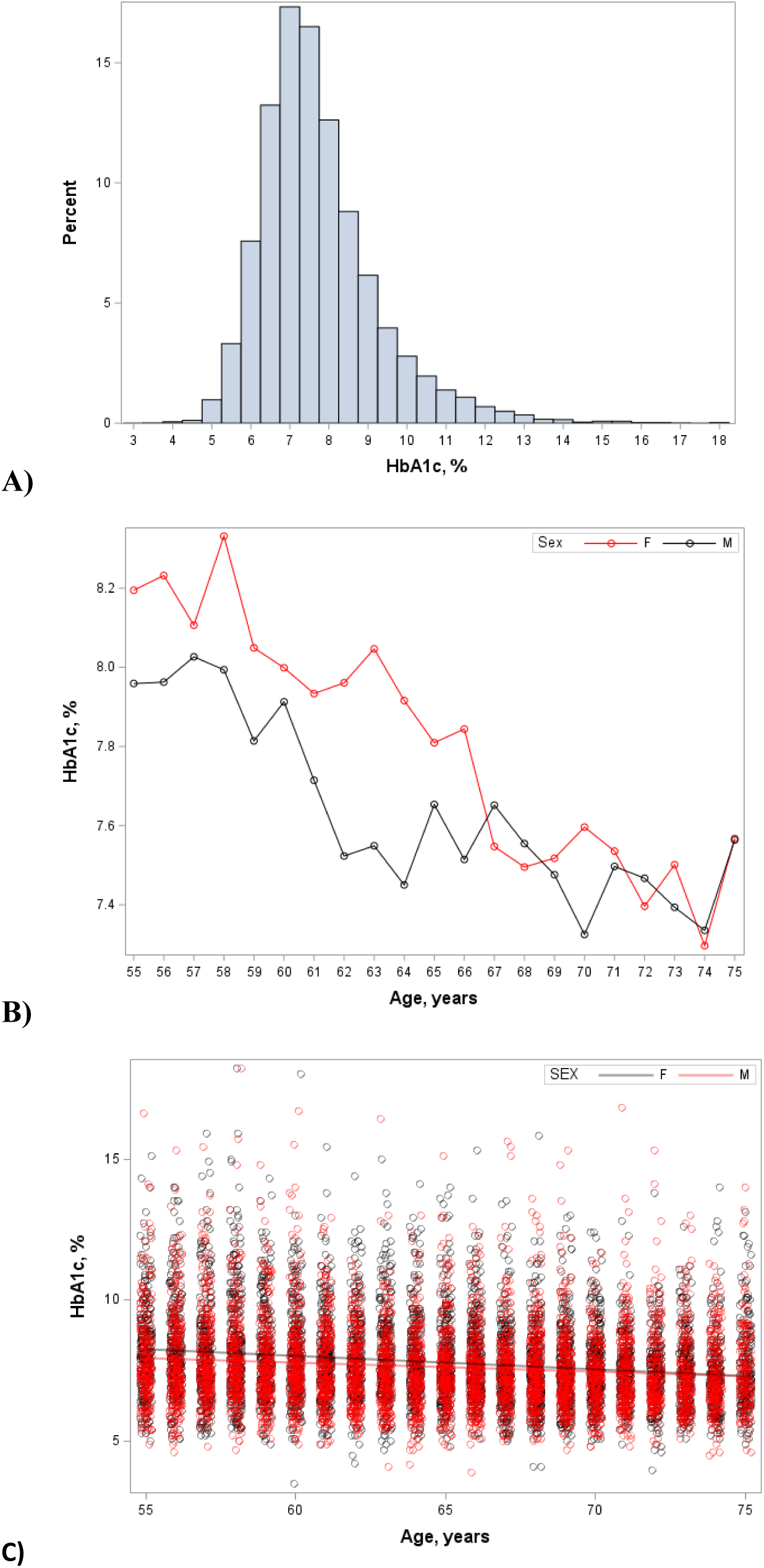
Hemoglobin A1c Distribution. Histogram of hemoglobin A1c frequency distribution for full cohort (**A**), mean hemoglobin A1c across age distribution by sex (**B**), and scatter plot of hemoglobin A1c with mean regression line by sex (**C**). Red line/circles identify females, and black line/circles identify males. Abbreviations: F, female; HbA1c, hemoglobin A1c (to convert to mmol/mol, 10.929*(HbA1c(%)-2.15); M, male.

**Table 1:** Cohort Descriptive Characteristics.

|  |  |
| --- | --- |
|  | Cohort with HbA1c data<br>(N= 12,088) |
| Age (years) | 64.7 ± 5.8 |
| Female gender, no./% | 6330 (52.4%) |
| Ethnicity, no./% | Hispanic: 331 (2.7%) |
|  | Non-Hispanic: 10,518 (87%) |
|  | Unknown: 1239 (10.3%) |
| Race, no./% | Asian: 58 (0.5%) |
|  | Black: 784 (6.5%) |
|  | Other: 419 (3.5%) |
|  | Unknown: 187 (1.5%) |
|  | White: 10640 (88%) |
| Marital status, no./% | Married: 6866 (56.8%) |
|  | Other: 266 (2.2%) |
|  | Single: 4889 (40.5%) |
|  | Unknown: 67 (0.5%) |
| Clinical Setting, no./% | Clinic: 827 (6.8%) |
|  | Diagnostic: 7229 (59.8%) |
|  | Home health/Rehabilitation: 25 (0.2%) |
|  | Observation: 1155 (9.5%) |
|  | Other: 1893 (15.7%) |
|  | Surgical: 890 (7.4%) |
|  | Infusion/hemodialysis: 69 (0.6%) |
| Insurance payor, no./% | Commercial: 602 (5%) |
|  | Managed Care: 3416 (28.3%) |
|  | Medicaid: 555 (4.6%) |
|  | Medicare: 7293 (60.3%) |
|  | Other: 114 (0.9%) |
|  | Uninsured: 108 (0.9%) |
| Hemoglobin A1c (%) | 7.7 ± 1.5 |
| Presence of ASCVD<br>diagnosis, no./% | 834 (6.9%) |

Abbreviations: ASCVD, atherosclerotic cardiovascular disease, HbA1c, to convert hemoglobin A1c % to mmol/mol, $10.929 \times (\text{HbA1c}(\%) - 2.15)$

In the multivariate regression model examining factors associated with HbA1c, several demographic characteristics were associated with HbA1c adjusting for all other variables (**Table 2**). Female sex conferred a 0.13% (95% CI: 0.08, 0.18, p < 0.0001) higher HbA1c on average. Each year of age was associated with a 0.04% lower HbA1c on average (95% CI: −0.04, −0.03, p<0.0001). Black race was associated with a 0.81% (95% CI: 0.70, 0.91, p < 0.0001) higher HbA1c compared to White race and 0.97% higher HbA1c compared to Asian race. Hispanic ethnicity was associated with a 0.26% higher HbA1c on average (95% CI: 0.09, 0.43, p = 0.002) compared to Non-Hispanic ethnicity. Medicaid insurance payor was associated with a 0.58% (95% CI: 0.41, 0.75, p < 0.0001) higher HbA1c compared to commercial insurance while Managed Care coverage was associated with a 0.18% lower HbA1c compared to commercial payor (effect plot examining HbA1c across insurance payors, Supplemental Figure S2). Examining clinical setting where the HbA1c was performed, testing at the time of an observational hospitalization was associated with a 0.41% higher HbA1c compared to clinic encounter (95% CI: 0.28, 0.55, p <0.0001). Pre-operative or outpatient surgical testing or other visit were both associated with lower HbA1c values compared to clinic setting. Single marital status, compared to married, was associated with a 0.15% higher HbA1c (95% CI: 0.10, 0.21, p < 0.0001). Presence of ASCVD diagnosis was not associated with a significantly higher HbA1c.

**Table 2:**
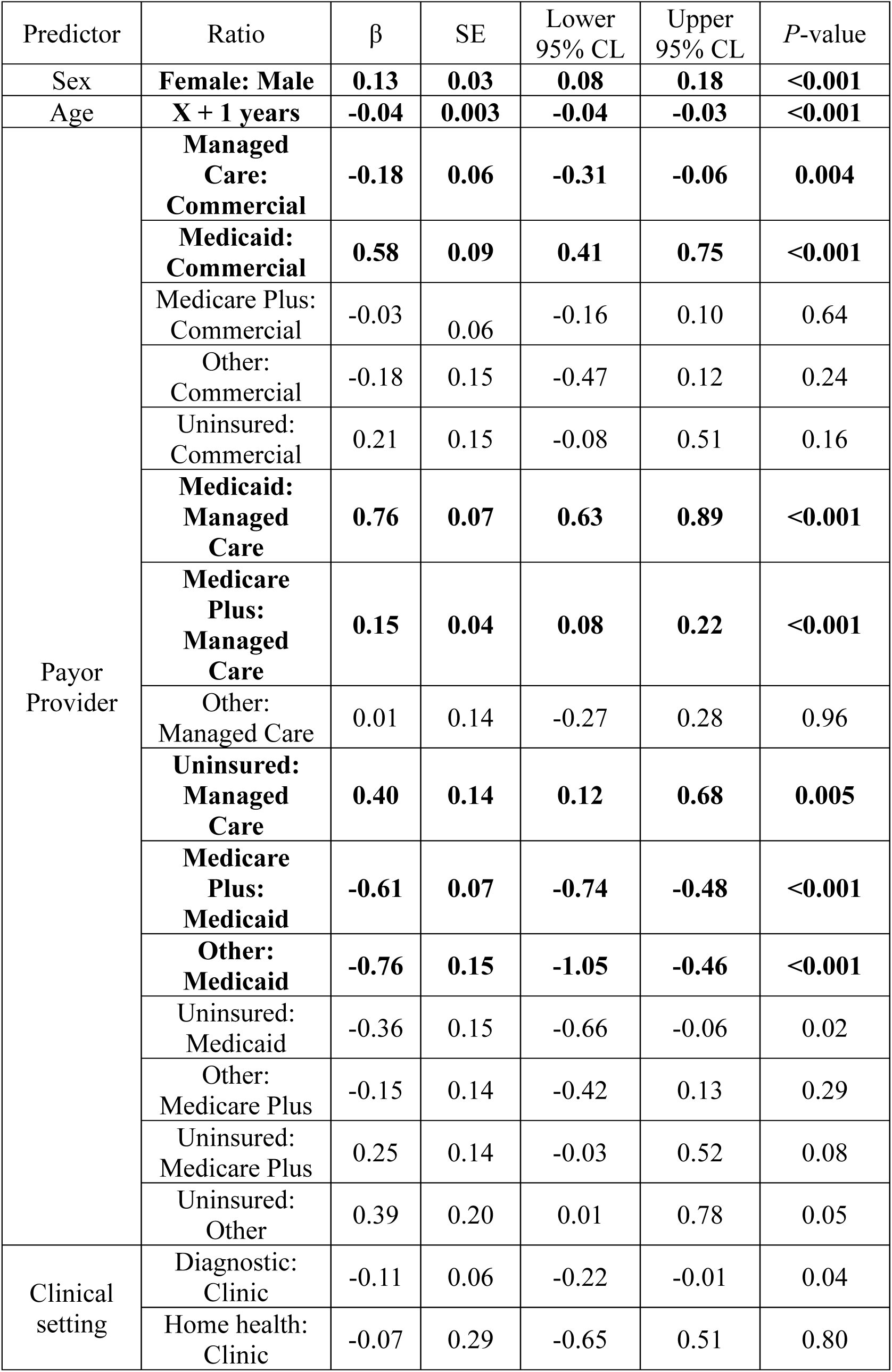
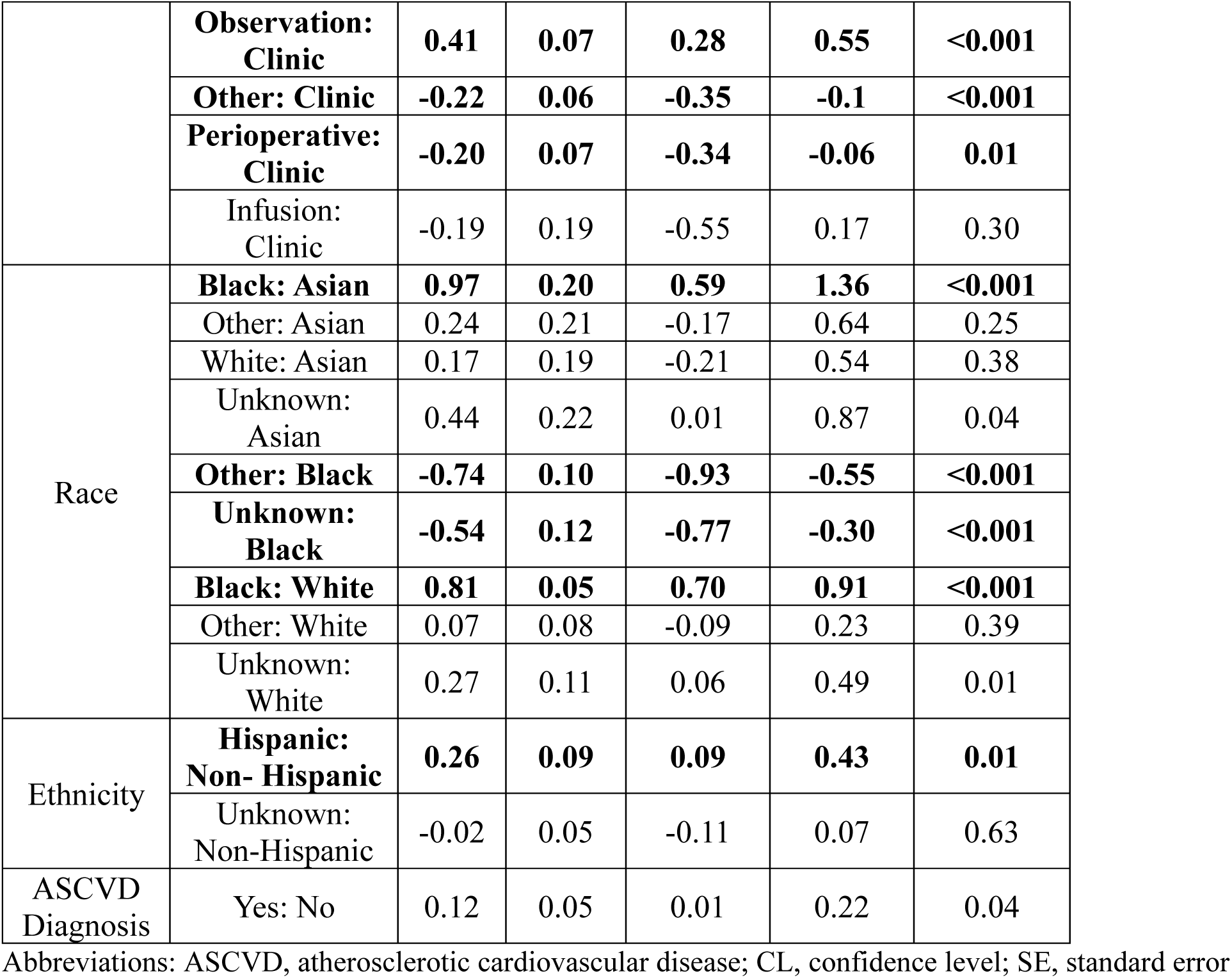
Multiple Variable Linear Regression Model for Predictors of Hemoglobin A1c.

We observed significant interactions for age and sex (p<0.0001), sex and race (p<0.0001). No significant interaction was present for ASCVD diagnosis and sex. Because of these interactions, a post-hoc stratified analysis by sex was performed to examine the influence of sex on model main effects (Supplemental Table S1). Females had a steeper HbA1c slope across age groups, indicating a greater reduction across ages and tendency towards HbA1c convergence at older ages (β=-0.05 for females, β=-0.02 for males, p<0.0001 for interaction). In racial and ethnic minority groups, men had higher HbA1c on average compared to women. On average, Black women had a 0.61% higher HbA1c while men had a 1.04% higher HbA1c compared to those identifying as White adjusting for all other variables. For women, ASCVD diagnosis was associated with a 0.18% higher HbA1c, while for men ASCVD diagnosis was associated with a 0.06% higher HbA1c, both were not significant.

In the multivariate logistic regression analysis examining demographic factors associated with HbA1c level above target and adjusting for all other variables, female sex, Black race, observational hospitalization compared to clinic encounter, single marital status, and presence of ASCVD diagnosis were each independently associated with HbA1c above target (**Figure 2**, Supplemental Tables S2 and S3). Females, compared to males, had a 16% higher odds of HbA1c above target adjusting for all other variables (adjusted OR 1.16, 95% CI: [1.07, 1.26]). Individuals with Black compared to White race had a 78% higher adjusted odds of HbA1c above target (adjusted odds ratio [OR] 1.78, 95% CI: 1.49, 2.12). On the other hand, individuals with Medicare and Managed Care insurance payors were more likely to have HbA1c at target. Younger age was associated with higher odds of HbA1c level above target.

**Figure 2:**
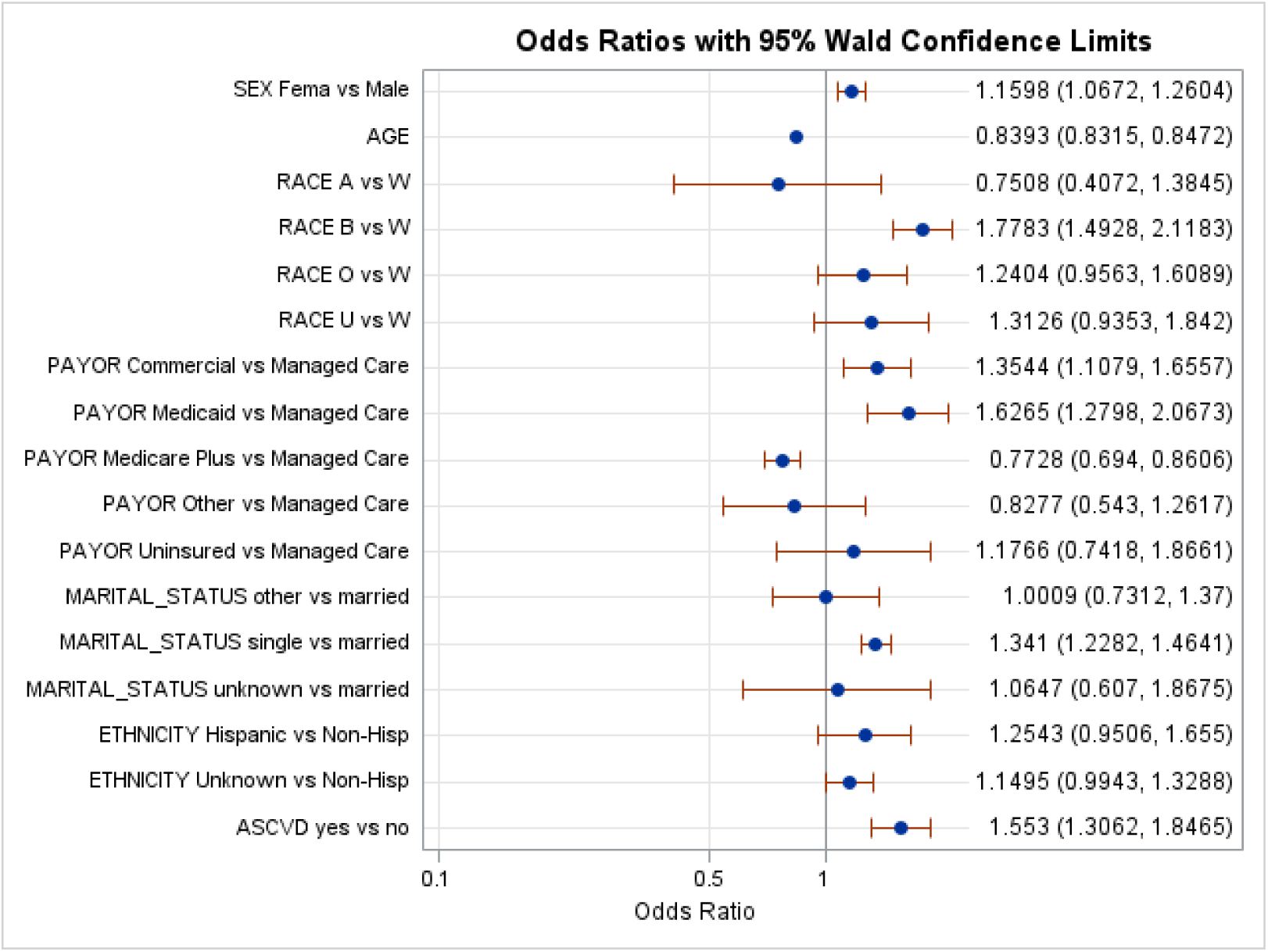
Adjusted Odds Ratio of Hemoglobin A1c Above Target. Abbreviations: A, Asian; ASCVD, atherosclerotic cardiovascular disease; B, black; O, other; U, unknown. Odds ratio plot of Hemoglobin A1c above target, adjusted for age, sex, race, ethnicity, insurance payor, testing location, marital status, and atherosclerotic cardiovascular disease.

## Conclusions

This study has several key findings. First, in this cohort with type 1 diabetes aged 55 to 75 years, women had a slightly higher HbA1c of 0.13% on average after adjusting for all other variables. While HbA1c was lower in advancing ages in both sexes, HbA1c trended down more steeply across ages in women. Few prior studies have included sufficient numbers of individuals over the age of 40 to detect a modest sex-based HbA1c difference in this age group; however, this data confirms results from several studies with older populations. In a large Swedish registry study (n = 16,367) with mean age 50.1 years, women had a significantly higher HbA1c adjusted for age and other socioeconomic factors like education level (11). In the 12^th^ year of EDIC, the observational cohort following the DCCT intervention study (n=1153), women had higher odds of HbA1c above target adjusting for age, diabetes duration, and activity level (12). Mean age and standard deviation in that study was 45 ± 7 years for women and 46 ± 7 years for men (12). Examining data from the Scottish Care Information Diabetes Collaboration, an EHR database including longitudinal HbA1c data from people with diabetes, women had a slightly higher HbA1c on average and increased risk of HbA1c above target which persisted across time, but this data was not grouped by age and sex simultaneously (13). A substantial proportion of that population, close to 50%, were 45 years or older. These results are also in line with data from the Canadian Diabetes National Repository. Examining longitudinal data from 296 patients with type 1 diabetes in the Canadian Diabetes National Repository, HbA1c decreased across the later lifespan and increased in women around the menopausal transition (16). In that study, tendency for glucose levels to increase around menopause was corroborated by community members with type 1 diabetes who participated in the study. They did not observe a significant age by sex interaction in the type 1 diabetes group, as was seen in the current investigation, and this may relate to limited sample size and wide age distribution. An interaction between sex and age was present for their larger type 2 diabetes cohort with women having a higher HbA1c in later age groups. The current cross-sectional study was not designed to investigate mechanisms for sex-based differences in HbA1c, but it underscores need for further investigation of women with type 1 diabetes around the menopause period to elucidate etiologies for these findings.

Our results contrast several large studies finding no HbA1c difference between sexes. In the U.S. Type 1 Diabetes Exchange Clinic Cohort, there was no sex-difference observed in HbA1c level or achievement of target HbA1c. Of the 8,674 individuals included, 2,810 were ages 50 years or older (14). Recent (2020–2022) data from the combined Prospective Diabetes Follow-up registry (DPV), in Germany, and the Société Francophone du Diabète–Cohorte Diabète de Type 1 cohort (SFDT1), in France, including 3,829 patients without cardiovascular disease history found no sex-difference in HbA1c or achieving target HbA1c levels (15). Mean age was 36-37 years in these cohorts. We hypothesize that the differences between these study findings and ours is the larger sample size of older patients, including the post-menopausal period.

The most concerning findings in this study relate to race and ethnicity-based differences in HbA1c levels which persisted after adjusting for insurance status, a factor which should partially moderate the impact of socioeconomic level. It is worth noting that multiple prior studies have observed a discordance between HbA1c and either mean CGM values or mean 7-point self-monitored blood glucose levels in Black individuals, with Black individuals having 0.33-0.4% higher HbA1c for a given mean glucose level than White individuals (20–22). On average, those identified as Black in this study had a 0.81% higher HbA1c compared to White and 0.97% higher HbA1c compared to Asian races adjusting for all other demographic variables. Even accounting for a HbA1c discordance, this is clinically significant. In fact, each longitudinal HbA1c increase of 1% is associated with a 26% increased CVD risk (10). Elevated kidney disease and CVD mortality are seen pervasively in the Black U.S. population. Although studies in type 1 diabetes are more limited, racial mortality outcome disparities are observed in the type 1 diabetes population, with Black patients having excess mortality risk (5,23). In a study of 18,767 adolescents and adult patients in the U.S. Type 1 Diabetes Exchange Registry cohort, participants identifying as Black were far more likely to have an above target HbA1c (adjusted OR 3.7, 95% CI: 1.9, 7.3)(24). Black race and Hispanic ethnicity remained associated with higher glycemia compared to Non-Hispanic White ethnicity even when restricting the analysis to the most socioeconomically deprived subgroup (24). Thus, factors outside of income, education, and health care access deprivation are contributing to these racial health disparities (24). In aggregate, the consistency of these findings are concerning for structural and implicit bias.

Higher HbA1c in the Hispanic U.S. population is also a consistent finding in the literature. In a meta-analysis of 11 studies including populations with type 1 and type 2 diabetes, the Hispanic population had an ∼0.5% higher HbA1c on average and more rapidly increasing trajectory compared to the non-Hispanic White population (25). A longitudinal analysis of 1313 youths with type 1 diabetes from the SEARCH for Diabetes in Youth cohort demonstrated that Hispanic youth had a 2.2 higher adjusted odds of being in the highest HbA1c group than non-Hispanic white youth (26). In a recent cross-sectional study of 12,035 patients from the Type 1 Diabetes Exchange Quality Improvement Collaborative, Hispanic and non-Hispanic Black populations were the groups with highest proportion of individuals with HbA1c above target (83.4% and 90.2% respectively)(27). We observed a 0.26% higher HbA1c in Hispanic compared to non-Hispanic patients in the multivariate regression model, but we did not observe a significantly increased risk of HbA1c above target in the multivariate logistic regression model. Adjustment for age moderated the impact of Hispanic ethnicity on HbA1c above target, suggesting this was a confounding factor. There were a limited number of Hispanic patients in this cohort, which may have reduced power to detect a difference.

The health disparities observed in the current study may relate in part to access to healthcare, insurance, diabetes technologies, differential prescribing habits related to implicit bias, and structural bias. U.S. young adults identifying as Hispanic and Non-Hispanic Black are less likely to utilize insulin pumps and continuous glucose monitors (28). Many barriers exist to health equity for people with type 1 diabetes under the current U.S. healthcare system and further interventions aimed at eradicating health disparities are needed. Offering continuous glucose monitor access within 1 month of diagnosis to all study participants with type 1 diabetes did in fact diminish racial and ethnic glycemic disparities in one recent study (29). More comprehensive reform including engaging community stakeholders, institutional examination of cardiometabolic data disaggregated by race/ethnicity/sex, cultural sensitivity and implicit bias training for medical providers, and a more diverse physician and research workforce are important future directions to tackle these structural inequities (30).

Our study also observed differences in HbA1c across insurance payors with Medicaid payor associated with highest HbA1c and Medicare and Managed Care ranking as lowest HbA1c (Supplemental Fig.2). In fact, individuals with Medicaid payor had a 0.76% higher HbA1c on average, adjusting for all other variables, compared to the Managed Care group. It is worth noting that HbA1c was not statistically different in uninsured (i.e. self-pay and indigent) compared to Medicaid payor groups in this study. Managed Care coverage conferred HbA1c advantage compared to Commercial and Medicaid insurances. These findings could relate to the preventative care model of Managed Care and Medicare insurances. Other recent studies also demonstrated disparities in glycemia when comparing U.S. insurance payors. In the SEARCH for Diabetes in Youth study, young adults without insurance demonstrated a higher mean HbA1c at 10.8% compared with 9.4% with public insurance (Medicare and Medicaid) and 8.7% with private insurance (31). Similar findings were noted in adults in the Type 1 Diabetes Exchange Quality Improvement Collaborative. In those participants, a majority of those achieving HbA1c values less than 7% had private insurance (59% private vs. 18% with public insurance) (27). Overall, odds of achieving HbA1c less than 7% was 1.47 times higher with private insurance compared with public (27). Individuals with generous insurance coverage have increased insulin pump and CGM utilization which translates to improved outcomes (32). Another potential explanation for these insurance-related disparities is lower socioeconomic status in groups with Medicaid insurance and young individuals with Medicare insurance. Furthermore, there are disparities in healthcare access based on medical practice insurance acceptance. Still, in adults with type 1 diabetes and type 2 diabetes at a major academic medical center which accepts Medicaid, patients with Medicaid payor had the highest rates of missed diabetes care appointments and higher HbA1c (33). Further root cause analysis is needed to understand causal factors for higher HbA1c in people using Medicaid insurance.

Finally, we observed that HbA1c was lower in older ages in both sexes. This was somewhat counterintuitive given less aggressive HbA1c targets beginning at age 65. This could relate to age of retirement for many patients, which allows additional time to focus on healthy habits such as physical activity, meal preparation, and optimizing diabetes management. We also considered whether the lower HbA1c in older age could relate to anemia. Anemia diagnosis by ICD-10 code was present in nearly 30% of the cohort and was slightly more common in adults over the age of 65. Adding anemia to the multivariate linear regression model did not change main effects of the other independent variables including age, suggesting that anemia was likely not a confounder for the lower HbA1c across age.

This study has several limitations. First, type 1 diabetes and ASCVD comorbidities were identified based on ICD-10 code. This diagnostic code was based on claims data but was not confirmed by additional validated EHR algorithms such as presence of hyperglycemia or majority of diabetes diagnosis codes for type 1 diabetes. This leads to lower specificity for the type 1 diabetes diagnosis and could misclassify patients with other types of diabetes or without diabetes as having type 1 diabetes. Further, ASCVD may have been under-reported if not a primary or secondary diagnosis. Second, we did not have access to data regarding diabetes technology use in this population which could be an important causal mediator in the disparities we observed. Third, most of the population in the Premier Healthcare Database with type 1 diabetes meeting eligibility criteria did not have HbA1c data. This could bias our results to reduce disparities in HbA1c for the most disadvantaged populations such as racial and ethnic minority groups if they did not have access to HbA1c assessment. In fact, the source type 1 diabetes population had a higher proportion of individuals identified as racial and ethnic minorities (8.4% Black, 5.5% Hispanic) compared to those with HbA1c data included in this cohort (6.5% Black, 2.7% Hispanic). We also did not have longitudinal data available to determine HbA1c change across time at the individual level. Nevertheless, this study has important strengths including a large population with type 1 diabetes in an older age group, accurate insurance payor data, comprehensive demographic data, and real world data which increases generalizability of these results.

In aggregate, in this cohort of patients with a type 1 diabetes diagnosis from the Premier Healthcare database, we observed a slightly higher HbA1c in women on average with lower HbA1c across older ages. Further investigation is needed to understand biological, and perhaps non-biological, factors which contribute to these trends. We also observed a clinically significant higher HbA1c in individuals identified as Black and individuals using Medicaid payor, adjusting for all other variables. Men in racial and ethnic minority groups were most profoundly impacted by higher HbA1c. Additional studies aimed at investigating glycemic changes around menopause and improving health equity are needed in the U.S. type 1 diabetes population.

## Data Availability

Data used in this study are available from Premier Healthcare Solutions, Inc. Restrictions apply to the availability of this data, which were used under license for the current study and therefore are non-transferable in accordance with a data use agreement with Premier Healthcare Solutions, Inc..

## Funding and Assistance

This work was supported by National Institutes of Health grant K23DK131327-01A1 (to K.M.L.). The content is solely the responsibility of the authors and does not necessarily represent the official views of the National Institutes of Health.

## Author contributions and Guarantor Statement

T.N.. and K.M.L. devised the hypothesis and wrote the first draft of the manuscript. K.M.L cleaned the data and performed statistical analysis. W.N. and G.R.L. contributed to statistical methods and data analysis and reviewed/edited the manuscript. S.A.B., M.C.S.V., and N.A.R. contributed to manuscript writing and reviewed/edited the manuscript. K.M.L. is the guarantor of this work, had full access to all the data in the study and takes responsibility for the integrity of the data and the accuracy of the data analysis.

## References

1. Soedamah-Muthu SS, Fuller JH, Mulnier HE, Raleigh VS, Lawrenson RA, Colhoun HM. High risk of cardiovascular disease in patients with type 1 diabetes in the U.K.: a cohort study using the general practice research database. Diabetes Care. 2006;29(4):798–804.

2. Tran-Duy A, Knight J, Palmer AJ, Petrie D, Lung TWC, Herman WH, Eliasson B, Svensson AM, Clarke PM. A Patient-Level Model to Estimate Lifetime Health Outcomes of Patients With Type 1 Diabetes. Diabetes Care. 2020;43(8):1741–1749.

3. Rawshani A, Svensson AM, Rosengren A, Eliasson B, Gudbjörnsdottir S. Impact of Socioeconomic Status on Cardiovascular Disease and Mortality in 24,947 Individuals With Type 1 Diabetes. Diabetes Care. 2015;38(8):1518–1527.

4. de Ferranti SD, de Boer IH, Fonseca V, Fox CS, Golden SH, Lavie CJ, Magge SN, Marx N, McGuire DK, Orchard TJ, Zinman B, Eckel RH. Type 1 diabetes mellitus and cardiovascular disease: a scientific statement from the American Heart Association and American Diabetes Association. Diabetes Care. 2014;37(10):2843–2863.

5. Roy M, Rendas-Baum R, Skurnick J. Mortality in African-Americans with Type 1 diabetes: The New Jersey 725. Diabet Med. 2006;23(6):698–706.

6. Laing SP, Swerdlow AJ, Slater SD, Burden AC, Morris A, Waugh NR, Gatling W, Bingley PJ, Patterson CC. Mortality from heart disease in a cohort of 23,000 patients with insulin-treated diabetes. Diabetologia. 2003;46(6):760–765.

7. Secrest AM, Becker DJ, Kelsey SF, Laporte RE, Orchard TJ. Cause-specific mortality trends in a large population-based cohort with long-standing childhood-onset type 1 diabetes. Diabetes. 2010;59(12):3216–3222.

8. Dart M, Patrie J, KM L. Demographic Predictors of Elevated LDL Cholesterol in Type 1 Diabetes: A Cross-Sectional Study. Clin Diabetes. 2024;10.2337/cd24-0.

9. Lind M, Svensson AM, Kosiborod M, Gudbjörnsdottir S, Pivodic A, Wedel H, Dahlqvist S, Clements M, Rosengren A. Glycemic control and excess mortality in type 1 diabetes. N Engl J Med. 2014;371(21):1972–1982.

10. Miller RG, Anderson SJ, Costacou T, Sekikawa A, Orchard TJ. Hemoglobin A1c Level and Cardiovascular Disease Incidence in Persons With Type 1 Diabetes: An Application of Joint Modeling of Longitudinal and Time-to-Event Data in the Pittsburgh Epidemiology of Diabetes Complications Study. Am J Epidemiol. 2018;187(7):1520–1529.

11. Willers C, Iderberg H, Axelsen M, Dahlström T, Julin B, Leksell J, Lindberg A, Lindgren P, Looström Muth K, Svensson AM, Lilja M. Sociodemographic determinants and health outcome variation in individuals with type 1 diabetes mellitus: A register-based study. PLoS One. 2018;13(6):e0199170.

12. Larkin ME, Backlund JY, Cleary P, Bayless M, Schaefer B, Canady J, Nathan DM, Group DCaCTEoDIaCDER. Disparity in management of diabetes and coronary heart disease risk factors by sex in DCCT/EDIC. Diabet Med. 2010;27(4):451–458.

13. Mair C, Wulaningsih W, Jeyam A, McGurnaghan S, Blackbourn L, Kennon B, Leese G, Lindsay R, McCrimmon RJ, McKnight J, Petrie JR, Sattar N, Wild SH, Conway N, Craigie I, Robertson K, Bath L, McKeigue PM, Colhoun HM, Group SDRNSE. Glycaemic control trends in people with type 1 diabetes in Scotland 2004-2016. Diabetologia. 2019;62(8):1375–1384.

14. Shah VN, Wu M, Polsky S, Snell-Bergeon JK, Sherr JL, Cengiz E, DiMeglio LA, Pop-Busui R, Mizokami-Stout K, Foster NC, Beck RW, Registry fTDEC. Gender differences in diabetes self-care in adults with type 1 diabetes: Findings from the T1D Exchange clinic registry. J Diabetes Complications. 2018;32(10):961–965.

15. Cosson E, Auzanneau M, Aguayo GA, Karges W, Riveline JP, Augstein P, Sablone L, Jehle P, Fagherazzi G, Holl RW, group DiatSs. Sex inequalities in cardiovascular risk factors and their management in primary prevention in adults living with type 1 diabetes in Germany and France: findings from DPV and SFDT1. Cardiovasc Diabetol. 2024;23(1):342.

16. Mousavi S, Tannenbaum Greenberg D, Ndjaboué R, Greiver M, Drescher O, Chipenda Dansokho S, Boutin D, Chouinard JM, Dostie S, Fenton R, Greenberg M, McGavock J, Najam A, Rekik M, Weisz T, Willison DJ, Durand A, Witteman HO, Study DACRQP. The Influence of Age, Sex, and Socioeconomic Status on Glycemic Control Among People With Type 1 and Type 2 Diabetes in Canada: Patient-Led Longitudinal Retrospective Cross-sectional Study With Multiple Time Points of Measurement. JMIR Diabetes. 2023;8:e35682.

17. Committee ADAPP. 13. Older Adults: Standards of Medical Care in Diabetes-2022. Diabetes Care. 2022;45(Suppl 1):S195–S207.

18. Association AD. 12. Older Adults:. Diabetes Care. 2020;43(Suppl 1):S152–S162.

19. Association AD. 12. Older Adults:. Diabetes Care. 2021;44(Suppl 1):S168–S179.

20. Karter AJ, Parker MM, Moffet HH, Gilliam LK. Racial and Ethnic Differences in the Association Between Mean Glucose and Hemoglobin A1c. Diabetes Technol Ther. 2023;25(10):697–704.

21. Bergenstal RM, Gal RL, Connor CG, Gubitosi-Klug R, Kruger D, Olson BA, Willi SM, Aleppo G, Weinstock RS, Wood J, Rickels M, DiMeglio LA, Bethin KE, Marcovina S, Tassopoulos A, Lee S, Massaro E, Bzdick S, Ichihara B, Markmann E, McGuigan P, Woerner S, Ecker M, Beck RW, Group TDERDS. Racial Differences in the Relationship of Glucose Concentrations and Hemoglobin A1c Levels. Ann Intern Med. 2017;167(2):95–102.

22. Wolffenbuttel BH, Herman WH, Gross JL, Dharmalingam M, Jiang HH, Hardin DS. Ethnic differences in glycemic markers in patients with type 2 diabetes. Diabetes Care. 2013;36(10):2931–2936.

23. Bosnyak Z, Nishimura R, Hagan Hughes M, Tajima N, Becker D, Tuomilehto J, Orchard TJ. Excess mortality in Black compared with White patients with Type 1 diabetes: an examination of underlying causes. Diabet Med. 2005;22(12):1636–1641.

24. Griggs S, Blanchette JE, Hickman RL, Magny-Normilus C, Baskin RG, Margevicius S, Hatipoglu B. Racial and Ethnic Cardiometabolic Risk Disparities in the Type 1 Diabetes Exchange Clinic Registry Cohort. Endocr Pract. 2022;28(12):1237–1243.

25. Kirk JK, Passmore LV, Bell RA, Narayan KM, D’Agostino RB, Arcury TA, Quandt SA. Disparities in A1C levels between Hispanic and non-Hispanic white adults with diabetes: a meta-analysis. Diabetes Care. 2008;31(2):240–246.

26. Kahkoska AR, Shay CM, Crandell J, Dabelea D, Imperatore G, Lawrence JM, Liese AD, Pihoker C, Reboussin BA, Agarwal S, Tooze JA, Wagenknecht LE, Zhong VW, Mayer-Davis EJ. Association of Race and Ethnicity With Glycemic Control and Hemoglobin A. JAMA Netw Open. 2018;1(5).

27. Akturk HK, Rompicherla S, Rioles N, Desimone M, Weinstock RS, Haw SJ, Ziemer DC, Dickinson JK, Agarwal S, Ebekozien O, Polsky S. Factors Associated With Improved A1C Among Adults With Type 1 Diabetes in the United States. Clin Diabetes. 2022;41(1):76–80.

28. Agarwal S, Schechter C, Gonzalez J, Long JA. Racial-Ethnic Disparities in Diabetes Technology use Among Young Adults with Type 1 Diabetes. Diabetes Technol Ther. 2021;23(4):306–313.

29. Addala A, Ding V, Zaharieva DP, Bishop FK, Adams AS, King AC, Johari R, Scheinker D, Hood KK, Desai M, Maahs DM, Prahalad P, Teamwork T, T.chnology, and Tight Control (4T) Study Group. Disparities in Hemoglobin A1c Levels in the First Year After Diagnosis Among Youths With Type 1 Diabetes Offered Continuous Glucose Monitoring. JAMA Netw Open. 2023;6(4):e238881.

30. Marshall A, Palokoff K. Dismissed: Tackling the Biases that Undermine Our Health Care. New York, NY: Kensington Publishing Corp.

31. Pihoker C, Braffett BH, Songer TJ, Herman WH, Tung M, Kuo S, Bellatorre A, Isganaitis E, Jensen ET, Divers J, Zhang P, Nathan DM, Drews K, Dabelea D, Zeitler PS, Group WCftSfDiYSGatTS. Diabetes Care Barriers, Use, and Health Outcomes in Younger Adults With Type 1 and Type 2 Diabetes. JAMA Netw Open. 2023;6(5):e2312147.

32. Everett EM, Wisk LE. Relationships Between Socioeconomic Status, Insurance Coverage for Diabetes Technology and Adverse Health in Patients With Type 1 Diabetes. J Diabetes Sci Technol. 2022;16(4):825–833.

33. Radhakrishnan R, Cade W, Bernal-Mizrachi E, Garg R. Medicaid insured persons with diabetes have increased proportion of missed appointments and high HbA1c. Am J Med Open. 2022;8:100022.

