## Supplemental Material for "Association of Sex and Other Demographic Characteristics with Hemoglobin A1c in Older Adults with Type 1 Diabetes: Cross-sectional analysis of the U.S. Premier Healthcare Database"

Online-Only Supplemental Material

Supplemental Figure S1: Cohort Flow Diagram

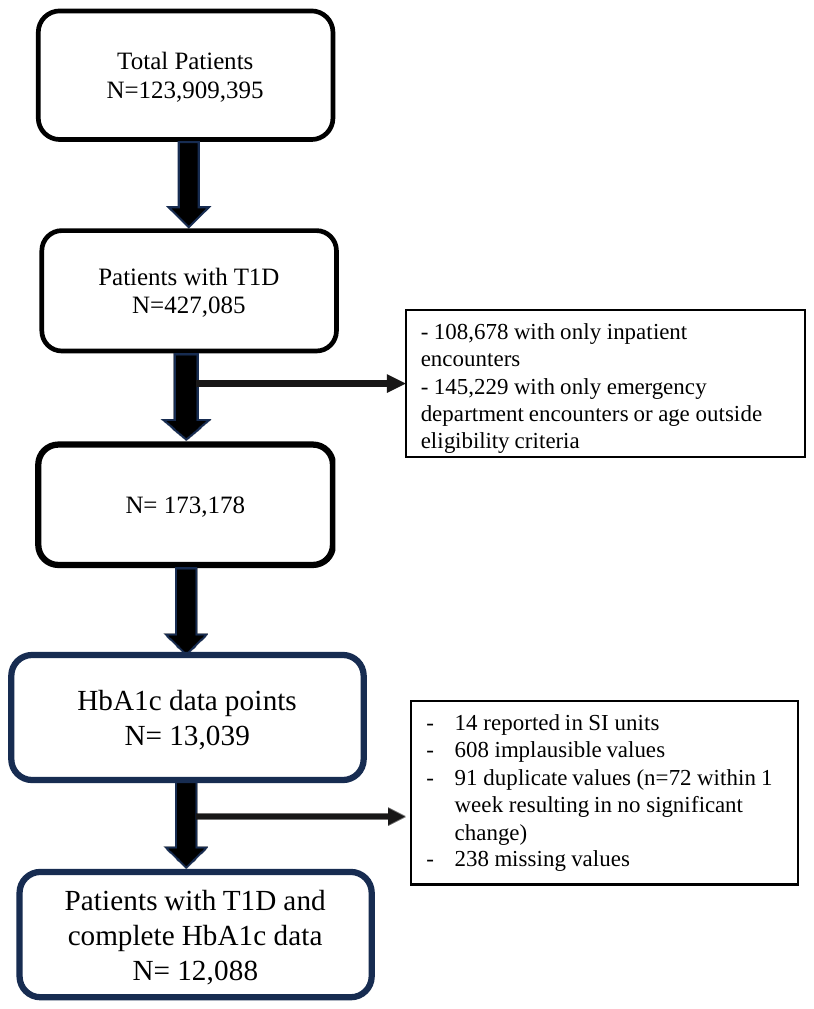

Supplemental Table S1: Multivariate linear regression for predictors of hemoglobin A1c, stratified by sex

|  |  | **Females** | | | | | **Males** | | | | |
| --- | --- | --- | --- | --- | --- | --- | --- | --- | --- | --- | --- |
| Predictor | Ratio | β | SE | Lower 95% CL | Upper 95%  CL | *p*-value | β | SE | Lower 95% CL | Upper 95%  CL | *p*-value |
| Age | X + 1 year | **-0.05** | **0.01** | **-0.06** | **-0.04** | **<0.001** | **-0.02** | **0.01** | **-0.03** | **-0.01** | **<0.001** |
| Race | Asian: White | -0.13 | 0.24 | -0.61 | 0.35 | 0.59 | -0.32 | 0.31 | -0.92 | 0.29 | 0.31 |
|  | **Black: White** | **0.61** | **0.08** | **0.47** | **0.76** | **<0.001** | **1.04** | **0.08** | **0.88** | **1.19** | **<0.001** |
|  | Other: White | 0.13 | 0.12 | -0.10 | 0.36 | 0.26 | 0.01 | 0.12 | -0.22 | 0.23 | 0.98 |
|  | Unknown: White | 0.25 | 0.17 | -0.08 | 0.57 | 0.15 | 0.32 | 0.14 | 0.04 | 0.60 | 0.03 |
|  | **Asian: Black** | **-0.75** | **0.25** | **-1.24** | **-0.25** | **0.01** | **-1.35** | 0.32 | -1.97 | -0.76 | **<0.001** |
|  | **Other:**  **Black** | **-0.48** | **0.14** | **-0.75** | **-0.22** | **<0.001** | **-1.03** | 0.14 | -1.30 | -0.76 | **<0.001** |
|  | Unknown: Black | -0.37 | 0.18 | -0.73 | -0.01 | 0.04 | **-0.72** | 0.16 | -1.03 | -0.40 | **<0.001** |
|  | Other:  Asian | 0.26 | 0.27 | -0.26 | 0.79 | 0.33 | 0.32 | 0.33 | -0.32 | 0.96 | 0.33 |
|  | Unknown: Asian | 0.38 | 0.29 | -0.20 | 0.95 | 0.20 | 0.63 | 0.34 | -0.03 | 1.30 | 0.06 |
|  | Unknown: Other | 0.11 | 0.20 | -0.27 | 0.50 | 0.56 | 0.32 | 0.18 | -0.03 | 0.66 | 0.07 |
| Ethnicity | **Hispanic:**  **Non-Hispanic** | 0.19 | 0.12 | -0.04 | 0.43 | 0.10 | **0.42** | 0.13 | 0.16 | 0.67 | **0.001** |
|  | Unknown:  Non-Hispanic | 0.03 | 0.07 | -0.11 | 0.16 | 0.71 | -0.07 | 0.06 | -0.20 | 0.05 | 0.25 |
|  | Unknown:  Hispanic | -0.17 | 0.13 | -0.42 | 0.08 | 0.19 | **-0.49** | 0.14 | -0.77 | -0.22 | **<0.001** |
| ASCVD | Yes: No | 0.18 | 0.08 | 0.01 | 0.34 | 0.03 | 0.06 | 0.07 | -0.08 | 0.20 | 0.4 |

Adjusted for all other variables listed in table, insurance payor, clinical setting, and marital status. Abbreviation: ASCVD, diagnosis code for atherosclerotic cardiovascular disease; CL, confidence limits

Supplemental Table S2: Unadjusted and adjusted odds ratio from Logistic Regression Model for Hemoglobin A1c above target (n = 12,088)

| Predictor | Ratio | Odds Ratio | Lower 95% CL | Upper 95% CL | *P*-value |
| --- | --- | --- | --- | --- | --- |
| Model 1: Unadjusted | | | | | |
| Sex | Female: Male | 1.13 | 1.05 | 1.21 | <0.001 |
| Race | Asian: White | 0.94 | 0.56 | 1.58 | 0.10 |
|  | Black: White | 2.07 | 1.78 | 2.40 | <0.001 |
|  | Other: White | 1.33 | 1.09 | 1.61 | 0.91 |
|  | Black: Asian | 2.20 | 1.29 | 3.77 | <0.001 |
|  | Other: Asian | 1.41 | 0.81 | 2.45 | 0.91 |
|  | Other: Black | 0.64 | 0.50 | 0.82 | 0.91 |
| Ethnicity | Hispanic: Non-Hispanic | 1.81 | 1.45 | 2.27 | <0.001 |
| Insurance Payor | Commercial: Managed Care | 1.29 | 1.07 | 1.56 | <0.01 |
|  | Medicaid: Managed Care | 2.44 | 1.94 | 3.06 | <0.001 |
|  | Medicare: Managed Care | 0.29 | 0.27 | 0.32 | <0.001 |
|  | Other: Managed Care | 0.77 | 0.53 | 1.13 | 0.11 |
|  | Uninsured: Managed Care | 1.48 | 0.96 | 2.27 | 0.04 |
| Model 2: Adjusted for age, sex | | | | | |
| Sex | Female: Male | 1.18 | 1.09 | 1.28 | <0.001 |
| Race | Asian: White | 0.73 | 0.40 | 1.34 | 0.05 |
|  | Black: White | 1.90 | 1.60 | 2.25 | <0.001 |
|  | Other: White | 1.32 | 1.06 | 1.66 | 0.36 |
|  | Black: Asian | 2.59 | 1.38 | 4.86 | <0.001 |
|  | Other: Asian | 1.81 | 0.95 | 3.45 | 0.36 |
|  | Other: Black | 0.70 | 0.53 | 0.92 | 0.36 |
| Ethnicity | Hispanic: Non-Hispanic | 1.44 | 1.11 | 1.85 | 0.04 |
| Insurance Payor | Commercial: Managed Care | 1.30 | 1.07 | 1.59 | 0.30 |
|  | Medicaid: Managed Care | 1.98 | 1.56 | 2.51 | <0.001 |
|  | Medicare: Managed Care | 0.85 | 0.766 | 0.94 | <0.001 |
|  | Other: Managed Care | 0.91 | 0.60 | 1.38 | 0.16 |
|  | Uninsured: Managed Care | 1.35 | 0.85 | 2.12 | 0.50 |
| Model 3: Adjusted for age, sex, race, ethnicity | | | | | |
| Sex | Female: Male | 1.18 | 1.09 | 1.28 | <0.001 |
| Race | Asian: White | 0.73 | 0.40 | 1.34 | 0.07 |
|  | Black: White | 1.93 | 1.62 | 2.29 | <0.001 |
|  | Other: White | 1.21 | 0.96 | 1.53 | 0.68 |
|  | Black: Asian | 2.64 | 1.41 | 4.95 | <0.001 |
|  | Other: Asian | 1.66 | 0.87 | 3.16 | 0.68 |
|  | Other: Black | 0.62 | 0.47 | 0.83 | 0.68 |
| Ethnicity | Hispanic: Non-Hispanic | 1.39 | 1.06 | 1.82 | 0.09 |
| Insurance Payor | Commercial: Managed Care | 1.29 | 1.06 | 1.57 | 0.18 |
|  | Medicaid: Managed Care | 1.84 | 1.45 | 2.33 | <0.001 |
|  | Medicare: Managed Care | 0.83 | 0.75 | 0.92 | <0.001 |
|  | Other: Managed Care | 0.88 | 0.58 | 1.33 | 0.15 |
|  | Uninsured: Managed Care | 1.24 | 0.78 | 1.96 | 0.66 |
| Model 4: Adjusted for age, sex, race, ethnicity, insurance payor | | | | | |
| Sex | Female: Male | 1.18 | 1.08 | 1.28 | <0.001 |
| Race | Asian: White | 0.72 | 0.39 | 1.32 | 0.07 |
|  | Black: White | 1.90 | 1.60 | 2.26 | <0.001 |
|  | Other: White | 1.18 | 0.93 | 1.50 | 0.73 |
|  | Black: Asian | 2.65 | 1.41 | 4.97 | <0.001 |
|  | Other: Asian | 1.65 | 0.86 | 3.16 | 0.73 |
|  | Other: Black | 0.62 | 0.47 | 0.83 | 0.73 |
| Ethnicity | Hispanic: Non-Hispanic | 1.37 | 1.05 | 1.80 | 0.10 |
| Insurance Payor | Commercial: Managed Care | 1.29 | 1.06 | 1.57 | 0.18 |
|  | Medicaid: Managed Care | 1.84 | 1.45 | 2.33 | <0.001 |
|  | Medicare: Managed Care | 0.83 | 0.75 | 0.92 | <0.001 |
|  | Other: Managed Care | 0.88 | 0.58 | 1.33 | 0.15 |
|  | Uninsured: Managed Care | 1.24 | 0.78 | 1.96 | 0.66 |
| Model 5: Adjusted for age, sex, race, ethnicity, insurance payor, marital status | | | | | |
| Sex | Female: Male | 1.15 | 1.06 | 1.25 | 0.001 |
| Race | Asian: White | 0.78 | 0.42 | 1.43 | 0.11 |
|  | Black: White | 1.85 | 1.56 | 2.20 | <0.001 |
|  | Other: White | 1.19 | 0.92 | 1.54 | 0.86 |
|  | Black: Asian | 2.38 | 1.27 | 4.47 | <0.001 |
|  | Other: Asian | 1.53 | 0.79 | 2.96 | 0.86 |
|  | Other: Black | 0.64 | 0.47 | 0.87 | 0.86 |
| Ethnicity | Hispanic: Non-Hispanic | 1.39 | 1.06 | 1.82 | 0.06 |
| Insurance Payor | Commercial: Managed Care | 1.30 | 1.07 | 1.59 | 0.08 |
|  | Medicaid: Managed Care | 1.68 | 1.33 | 2.14 | <0.001 |
|  | Medicare: Managed Care | 0.78 | 0.70 | 0.87 | <0.001 |
|  | Other: Managed Care | 0.87 | 0.57 | 1.32 | 0.19 |
|  | Uninsured: Managed Care | 1.18 | 0.74 | 1.87 | 0.71 |
| Model 6: Age, sex, race, ethnicity, payor, marital status, clinical location | | | | | |
| Sex | Female: Male | 1.15 | 1.06 | 1.25 | <0.001 |
| Race | Asian: White | 0.75 | 0.41 | 1.39 | 0.08 |
|  | Black: White | 1.78 | 1.50 | 2.12 | <0.001 |
|  | Other: White | 1.26 | 0.97 | 1.63 | 0.58 |
|  | Black: Asian | 2.36 | 1.25 | 4.45 | <0.001 |
|  | Other: Asian | 1.66 | 0.86 | 3.22 | 0.58 |
|  | Other: Black | 0.70 | 0.52 | 0.96 | 0.58 |
| Ethnicity | Hispanic: Non-Hispanic | 1.25 | 0.95 | 1.65 | 0.26 |
| Insurance Payor | Commercial: Managed Care | 1.36 | 1.12 | 1.67 | 0.02 |
|  | Medicaid: Managed Care | 1.63 | 1.29 | 2.08 | <0.001 |
|  | Medicare: Managed Care | 0.78 | 0.70 | 0.87 | <0.001 |
|  | Other: Managed Care | 0.84 | 0.55 | 1.28 | 0.15 |

Supplemental Table S3: Type III Wald test ANOVA summary derived from multiple logistic regression model for Hemoglobin A1c above target (n=12,088)

| Predictor | Degrees of Freedom | Wald Type III Chi-square Statistic | P-value |
| --- | --- | --- | --- |
| Age | 1 | 1357.8 | **<0.001** |
| Sex | 1 | 12.2 | **<0.001** |
| Race | 4 | 46.4 | **<0.001** |
| Insurance Payor | 5 | 67.2 | **<0.001** |
| Ethnicity | 2 | 5.5 | 0.06 |
| Marital status | 3 | 43.5 | <0.001 |
| Clinical setting | 6 | 33.0 | <0.001 |
| ASCVD | 1 | 24.9 | <0.001 |
| TOTAL | **23** | **2459.3** | **<0.001** |
|  | *Model C-statistic* | *0.789* |  |
| **Hosmer-Lemeshow Goodness of Fit Test** | | | |
| Test | Degrees of Freedom | Chi-square Statistic | P-value |
| Goodness of Fit | 8 | 320.4 | <0.001 |

Abbreviations: ASCVD, atherosclerotic cardiovascular disease diagnosis

Supplemental Figure S2: Effect plot for hemoglobin A1c by insurance payor, from multivariate linear regression model

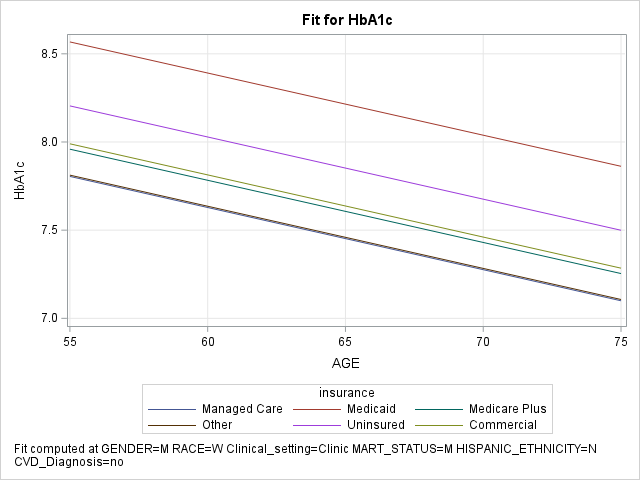

Effect plot computed at male sex, clinic location, married marital status, Non-Hispanic ethnicity, no atherosclerotic cardiovascular disease diagnosis, and adjusted for age.
